# Experiences of physiotherapists working with adults living with Long COVID in Canada: a qualitative study

**DOI:** 10.1101/2024.03.10.24304061

**Authors:** Caleb Kim, Chantal Lin, Michelle Wong, Shahd Al Hamour Al Jarad, Amy Gao, Nicole Kaufman, Kiera McDuff, Darren A. Brown, Saul Cobbing, Alyssa Minor, Soo Chan Carusone, Kelly K. O’Brien

## Abstract

**Objectives:** To explore experiences of physiotherapists working with adults living with Long COVID in Canada.

**Design:** Cross-sectional descriptive qualitative study involving online semi-structured interviews.

**Participants:** We recruited physiotherapists in Canada who self-identified as having clinically treated one or more adults living with Long COVID in the past year.

**Data collection:** Using an interview guide, we inquired about physiotherapists’ knowledge of Long COVID, assessment and treatment experiences, perspectives on physiotherapists’ roles, contextual and implementation factors influencing rehabilitative outcomes, and their recommendations for Long COVID rehabilitation. Interviews were audio-recorded, transcribed verbatim, and analyzed using a group-based thematic analytical approach. We administered a demographic questionnaire to describe sample characteristics.

**Results:** Thirteen physiotherapists from five provinces participated; most were women (n=8;62%) and practised in urban settings (n=11;85%). Participants reported variable amounts of knowledge of existing guidelines and experiences working with adults living with Long COVID in the past year. Physiotherapists characterized their experiences working with adults living with Long COVID as a dynamic process involving: 1) a disruption to the profession (encountering a new patient population and pivoting to new models of care delivery), followed by 2) a cyclical process of learning curves and evolving roles of physiotherapists working with persons living with Long COVID (navigating uncertainty, keeping up with rapidly-emerging evidence, trial and error, adapting mindset and rehabilitative approaches, and growing prominence of roles as advocate and collaborator). Participants recommended the need for education and training, active and open-minded listening with patients, interdisciplinary models of care, and organizational- and system-level improvements to foster access to care.

**Conclusions:** Physiotherapists’ experiences involved a disruption to the profession followed by a dynamic process of learning curves and evolving roles in Long COVID rehabilitation. Not all participants demonstrated an in-depth understanding of existing Long COVID rehabilitation guidelines. Results may help inform physiotherapy education in Long COVID rehabilitation.

**STRENGTHS AND LIMITATIONS OF THIS STUDY:**

- To our knowledge, this is one of the first qualitative studies to explore experiences of physiotherapists working with patients living with Long COVID in Canada.
- Our qualitative approach, involving online semi-structured interviews, enabled an in-depth exploration into Canadian physiotherapists’ perceived roles in Long COVID care, experiences with assessment and treatment, knowledge acquisition, and facilitators and barriers to delivery of rehabilitation services.
- Our team-based analytical approach and partnership with physiotherapists living with Long COVID as part of our process provided valuable collaboration, guidance, and advice for refining the interview guide and demographic questionnaire and fostering student researcher interview skills to increase the quality of the study.
- The diversity of participants’ characteristics working in different practice settings across five Canadian provinces and the variability in the number of individuals with Long COVID treated by participants were strengths of the study. However, as most participants practised in urban settings in Canada, transferability to other geographical contexts including rural settings and other countries may be limited, especially those with larger differences in healthcare systems.

## INTRODUCTION

The long-term sequelae of infection with the SARS-CoV-2 virus, otherwise known as Long COVID, is a global health concern. Long COVID, a term first coined by patients with lived experiences,(1,2) is defined as a condition: “occur[ing] in individuals with a history of probable or confirmed SARS CoV-2 infection, usually three months from the onset of COVID-19 with symptoms that last for at least two months and cannot be explained by an alternative diagnosis“(3). Globally, an estimated 144.7 million persons were living with Long COVID in 2020 and 2021, with approximately one in nine (12%) or 3.5 million Canadians experiencing long-term sequelae of COVID-19 in Canada as of 2023 (4,5).

Long COVID is a multisystemic condition that can manifest as multiple symptoms experienced as episodic, relapsing, and remitting over time (6,7). People living with Long COVID can experience one or many of over 200 symptoms affecting multiple organ systems (8–11). Post-exertional symptom exacerbation or post-exertional malaise (PESE or PEM), worsening or relapse of symptoms following minimal exertion such as physical or cognitive activity, is a common and debilitating health challenge experienced among many persons living with Long COVID (10,12–14). Other symptoms such as fatigue, persistent cough, dyspnea, and cognitive impairments were prevalent among persons living with Long COVID who were hospitalized or non-hospitalized with COVID-19 (9,10,12–17). In Canada, commonly reported Long COVID symptoms included fatigue (72%), cough (39%), shortness of breath (38%), and brain fog (33%) (18), which may negatively impact health-related quality of life of people living with Long COVID (15,16).

Together, the experiences among persons living with Long COVID can be characterized as disability, defined as any physical, cognitive, mental, and emotional health challenges, difficulties with daily activities, social inclusion, and uncertainty about future health, that may fluctuate on a daily basis or over the long term (19–21). People living with Long COVID described episodic fluctuations in health and disability as “ups and downs”, “relapsing and remitting”, or a “rollercoaster ride” (6,7), intersecting with uncertainty and worrying about the future. The severity, timing, and impact of episodes of disability may be unpredictable throughout the day, as well as over time. Episodic disability experienced by people with Long COVID has further societal implications such as challenges returning to work for those who have left the workforce, and financial and housing insecurity (22–28).

Safe rehabilitation, focused on symptom stabilization and avoiding exacerbation, has a potential role in addressing the multidimensional nature of disability experienced by people living with Long COVID (16,17,29). The World Health Organization (WHO) COVID-19 Clinical Management: Living Guideline (30) highlights the importance of multidisciplinary, coordinated, person-centred, and safe rehabilitation with persons living with Long COVID (6,7,31,32). Specifically, physiotherapists (physical therapists), may help to mitigate disability and prevent symptoms associated with Long COVID such as dyspnea, fatigue, and pain, and guide workplace accommodations (7,17,31–38). It is critical for physiotherapists to screen for complications from COVID-19 before rehabilitation and on an ongoing basis, including PESE, cardiac impairment, exertional oxygen desaturation, and autonomic dysfunction. Rehabilitation interventions must be modified for postural orthostatic tachycardia syndrome (POTS) and PESE to ensure safe approaches to assessment and treatment (30,39). Despite existing guidelines for safe rehabilitation in the context of Long COVID (30), the knowledge of physiotherapists, perceptions of their roles, and experiences in clinical practice working with persons living with Long COVID are unknown. We aimed to explore the experiences of physiotherapists working with adults living with Long COVID in Canada.

## METHODS

### Study Design

We conducted a cross-sectional qualitative descriptive study involving online semi-structured interviews with physiotherapists in Canada who worked clinically in the past year with one or more adults living with Long COVID (40).

### Patient and Public Involvement

This study involved a collaboration with Long COVID Physio, an international association of Physiotherapists living with Long COVID, providing peer support, education, and advocacy (41). Our team included current and future physiotherapists, researchers, and two persons with lived or living experiences of Long COVID. We consulted two physiotherapists living with Long COVID for their guidance when developing and refining the interview guide and the demographic questionnaire.

### Participants and Recruitment

We recruited English-speaking physiotherapists registered to practice in Canada who self-identified as having clinical experiences working with adults living with Long COVID in the past year. We defined clinical experiences as, but not limited to: practising in a Long COVID-designated setting (e.g. seeing only patients with Long COVID) and/or involving assessment and treatment of ≥1 adult(s) living with Long COVID in the past year as part of a mixed caseload.

We identified potential participants through professional networks in Long COVID care, word of mouth, and the use of search engines for Long COVID physiotherapy clinics. We emailed potential participants with an outline of the study, reason for study invitation, eligibility, information letter and consent form, and requested their participation. We asked participants to forward our recruitment email to fellow physiotherapists who met the inclusion criteria.

We used purposive and snowball sampling to recruit participants with diversity in clinical experience (estimated number of patients living with Long COVID treated in the past year), geographical location (urban versus rural), and practice setting (working in a designated Long COVID setting versus not) (42,43).

### Data Collection

#### Interviews

We conducted 2:1 (researcher:participant) semi-structured interviews online via Zoom (44) during which one researcher facilitated the discussion, and the other assisted with documenting field notes and providing reflections on the interview (45). We asked participants about their experiences and perspectives on the role of physiotherapists working with persons living with Long COVID - specifically, their knowledge and expertise of the condition, clinical practice setting, assessments, treatments, and patient-reported and objective outcome measures, as well as contextual and implementation factors that impact Long COVID rehabilitation. Additionally, we asked about their knowledge of guidelines for Long COVID rehabilitation, as well as recommendations they had for future rehabilitation programs and physiotherapists working with people with Long COVID (Supplemental File 1).

We conducted five mock interviews among the research team and refined the interview guide three times throughout to enhance clarity, maximize our ability to elicit participant responses in subsequent interviews, and enhance accessibility.

#### Demographic Questionnaire

We administered a two-part demographic questionnaire with interviewers entering responses into the questionnaires using Microsoft Word (46) to describe characteristics of study participants. Prior to the interview, we asked questions about professional characteristics, including clinical experiences in Long COVID care, workplace, years of practice, setting, location, extent of their work with adults living with Long COVID, and whether participants had personal lived experiences with Long COVID. After the interview we asked about personal characteristics such as age, sex, gender, and race/ethnicity (Supplemental File 2). Participants received a $40 CAD electronic gift card as a token of appreciation for their participation in the study.

### Ethics and Consent

This study protocol was approved by the University of Toronto Health Sciences Research Ethics Board (Protocol #00043692) (Supplemental File 3). All participants provided verbal consent to participate in the study, which was recorded by the interviewers by signing and dating the consent form (Supplemental File 4).

### Data Analysis

Interviews were audio-recorded, then transcribed verbatim and reviewed for accuracy by the primary interviewer (47). Next, we performed a qualitative descriptive analysis of the transcripts and field notes. Initially, all team members independently read and coded the same three transcripts line-by-line to create codes that operationalized ‘experiences’ working with adults living with Long COVID. We then grouped common responses and terms into categories to describe the experiences of physiotherapists. After coding a fourth transcript, we developed a preliminary coding scheme that included codes related to concepts characterizing experiences working with adults living with Long COVID. Dyads of team members independently coded each of the remaining transcripts and discussed the coding process. We then met as a full team two additional times to refine the coding scheme. Participant summaries were developed for each interview that included participant demographic information and a summary of overall impressions and highlights from the interview. We used Microsoft Excel (48) to organize our data according to codes and broader categories.

We analyzed the demographic questionnaire responses by calculating frequencies (%) for categorical variables, and median and 25-75th percentiles for continuous variables.

### Sample Size

We aimed to recruit 12-15 participants. Authors of a qualitative descriptive study who investigated community healthcare workers’ experiences during the COVID-19 pandemic (49) had a similar sample size (n=15) and were able to address their study objectives. Similarly, authors of two qualitative descriptive interview-based studies with comparable sample sizes (n=11, n=15) achieved their objectives, which were to understand the experiences of physiotherapists who had implemented a chronic pain self-management programme in primary health care (50) and to identify factors to consider for developing and implementing a community-based exercise program for people living with HIV from the perspectives of people living with HIV, rehabilitation professionals, and recreation providers (51).

## RESULTS

Out of the 54 individuals who expressed interest in the study, 13 (24%) physiotherapists met the inclusion criteria and agreed to participate in the study. Interviews were held between February and June 2023 and had a median (min-max) duration of 56 (45–70) minutes.

### Characteristics of Participants

#### Personal Characteristics

Thirteen physiotherapists in Canada participated in an interview. The median age of participants was 41 years (25th, 75th percentile: 34, 50), and the majority identified as women (n=8; 62%). Most participants practiced in an urban setting (n=11; 85%), representing five Canadian provinces including Ontario (n=5; 38%), Alberta (n=4; 31%), British Columbia (n=2; 15%), Quebec (n=1; 8%), and Nova Scotia (n=1; 8%). Two participants (15%) reported having lived experiences of Long COVID. *Clinical Characteristics* In the past year, five (38%) physiotherapists treated between 1 to 25 patients living with Long COVID, four (31%) treated between 26 to 100, and four (31%) treated over 100. Practice settings in which physiotherapists worked with adults living with Long COVID included outpatient or inpatient rehabilitation hospitals (n=5; 38%), private practice clinics (n=4; 31%), and publicly-funded outpatient clinics within a general or community hospital (n=4; 31%). The majority of physiotherapists (n=12; 92%) delivered a proportion of their caseload virtually, with seven (54%) delivering ≥ 50% of their care virtually to patients with Long COVID. Additionally, three (23%) participants reported experiences working with children or youth (<18 years old) living with Long COVID. See Table 1 for additional participant characteristics.

**Table 1.**
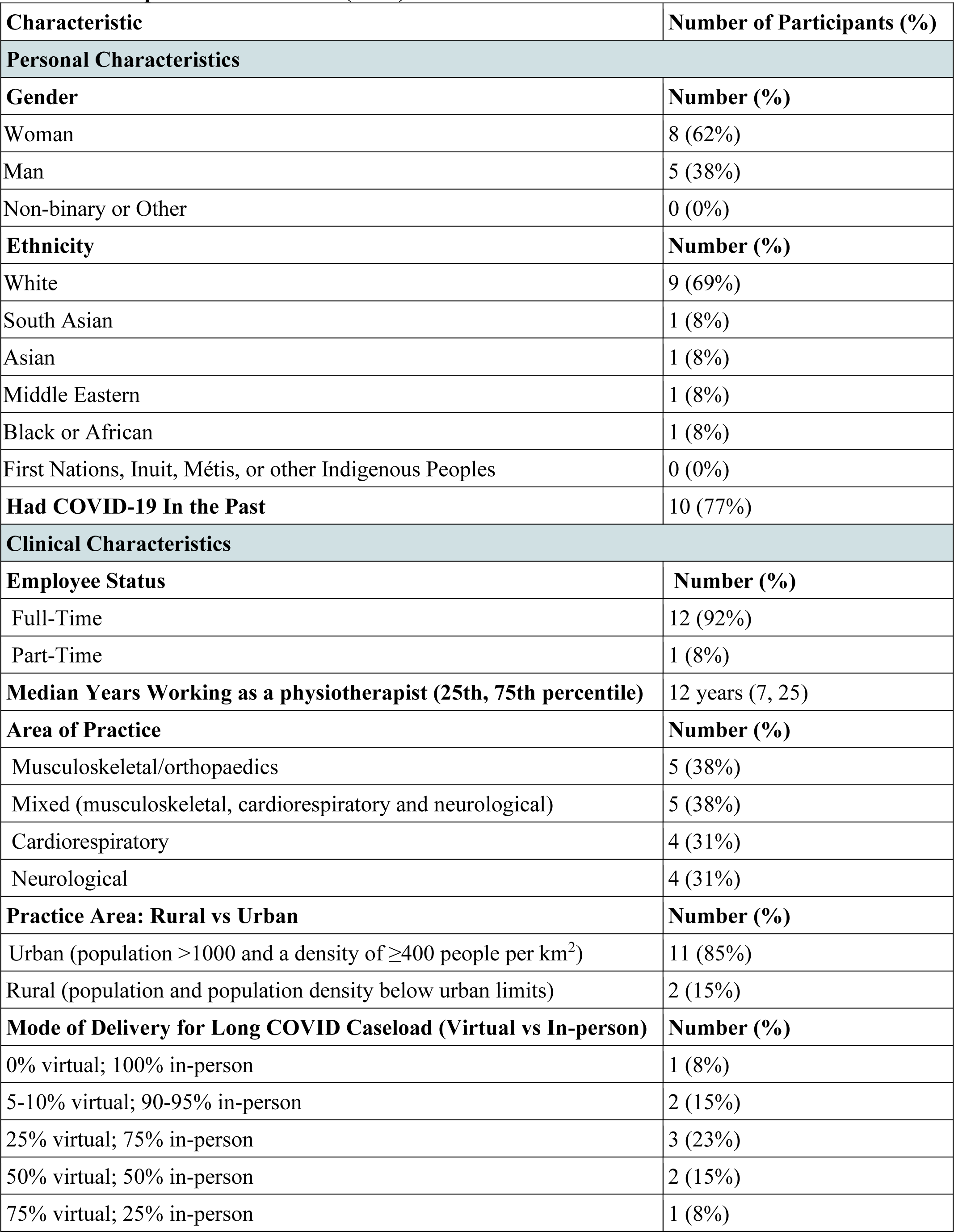
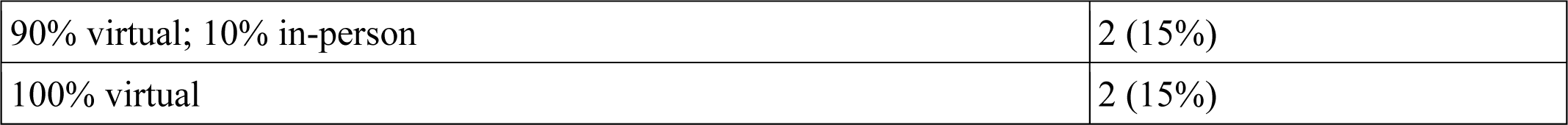
Participant Characteristics (n=13)

#### Physiotherapists with Lived Experiences with Long COVID

Two physiotherapists reported having lived experiences with Long COVID. They described how their experiences impacted their practice within Long COVID care, approaching it with “*a lot more empathy*” (P5) and helping to “*[connect] with a patient level*” (P10). They “*better understood what [patients] were referring to when they said [their body] didn’t feel normal”* (P5) while acknowledging that experiences are individual.

#### Experiences of Physiotherapists in Long COVID Rehabilitation

Experiences of physiotherapists in Canada working with adults with Long COVID in this sample were characterized by a dynamic process involving two components: 1) a disruption to the profession and models of physiotherapy care delivery prompted by the COVID-19 pandemic and a new population of people living with Long COVID, followed by 2) a cyclical journey involving learning curves and evolving roles of physiotherapists working with people living with Long COVID (Figure 1). We describe these components, and categories that comprise each, with supporting quotations below.

**Figure 1.**
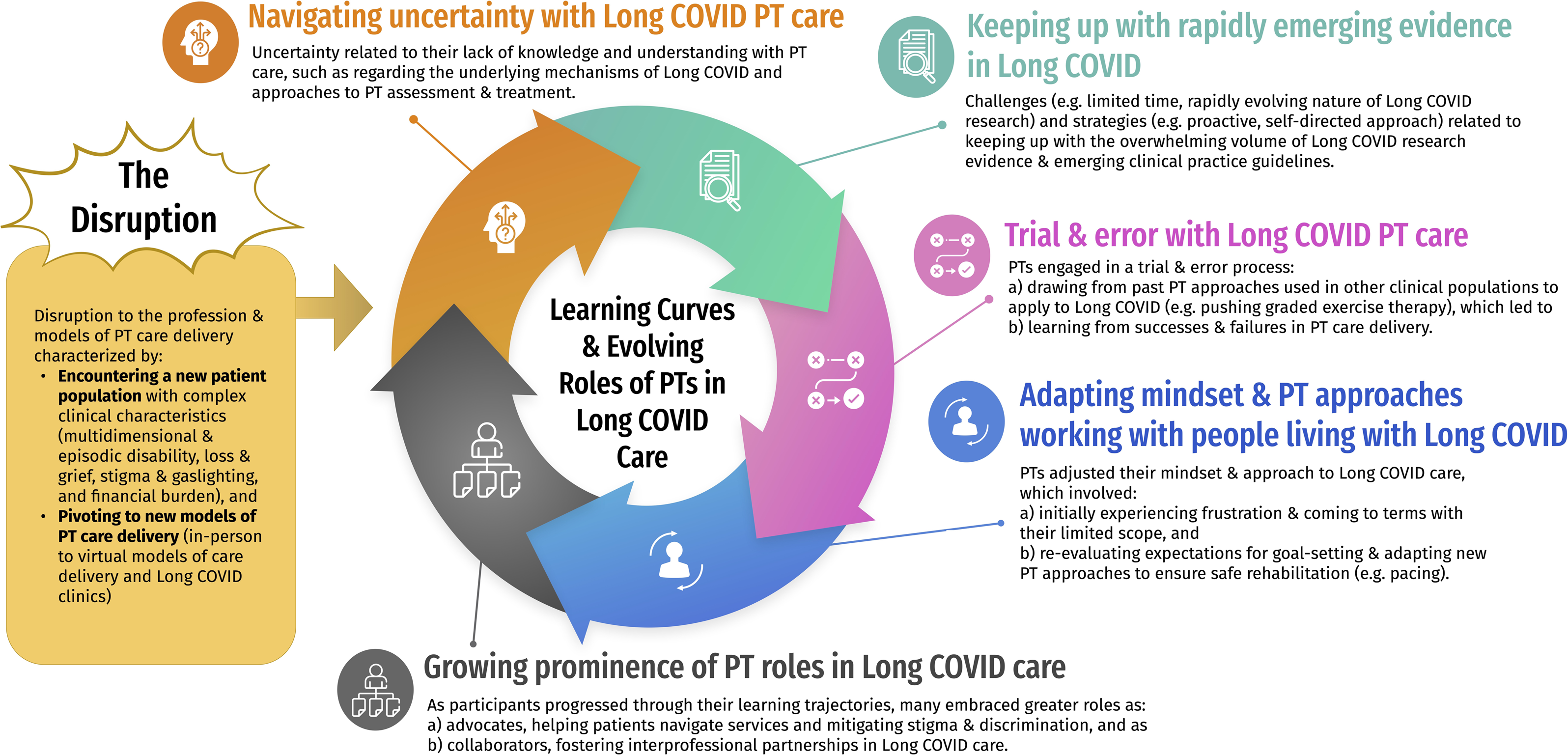
Experiences of Physiotherapists Working with Adults Living With Long COVID

### The Disruption

Participants described the onset of the COVID-19 pandemic and encountering the first patients with Long COVID in their clinical practice as a momentous event, requiring a shift in the way they approached education and clinical care delivery, and the challenges required to adapt to adequately meet the needs of persons living with Long COVID. Physiotherapists described i) encountering a new patient population with complex clinical characteristics (multidimensional and episodic disability; loss and grief; stigma and testimonial injustice (gaslighting); financial burden), and ii) pivoting to new models of physiotherapy care delivery (in-person to virtual models of care delivery; Long COVID clinics).

#### Encountering a New Patient Population

Participants discussed the challenges associated with an increasing caseload of patients presenting with Long COVID. They reported that health challenges experienced by patients with Long COVID included a) physical, cognitive, and emotional symptoms affecting daily life and social participation, situated within b) contextual factors such as stigma and testimonial injustice (gaslighting) that worsened episodes of disability. Many physiotherapists were unfamiliar with the chronic and episodic nature of Long COVID. Participants identified the variable and chronic nature of Long COVID symptoms as unique relative to more common conditions they encountered in their clinical practices. Long COVID was associated with fluctuating levels of function, without symptom resolution in response to treatment. This physiotherapist described their lack of understanding of Long COVID, including confusion about its definition, its complexity, and the multidimensional health-related challenges of the condition:

> I don’t think I had a full understanding of what it was. I feel like the definition changed a lot… what encompassed Long COVID was different than…what it is now…So I would say that the symptoms that I’m now including into my personal definition of Long COVID has increased. (P11)

Physiotherapist participants described encountering both familiar and unfamiliar symptoms of Long COVID, such as “*fatigue*”, “*PEM/PESE*”, *“exercise intolerance*”, “*dysautonomia*”, “*POTS [Postural Orthostatic Tachycardia Syndrome]*”, “*brain fog*” [cognitive dysfunction], “*chest tightness*” and “*high heart rate variability*”. Physiotherapists were learning about these symptoms from their patients with Long COVID. One participant articulated a patient’s description of their fatigue as *“a wave of fatigue and you just had to let it rush through your body and let it pass before you could even start to do anything after that.”* (P5) Additionally, participants described learning from concerns their patients articulated about the potential impacts of re-infection and the unpredictability of the condition: “it was something that nobody understood so nobody could give them specific answers…It’s very scary when you don’t understand what’s happening in your body.” (P3)

Participants described learning from their patients about “*illness adjustment*” (P9), the impacts of loss of function and independence, and the journey of acceptance and redefinition of self-identity and community roles:

> [Patients with Long COVID]… because their identity was tied in with being able to do everything around the house, and to be that person that everyone relies on…now [they] almost have to kind of redefine who they are as a person, whether it be your family or their community, or as a provider… (P5)

Many participants described stigma and testimonial injustice (gaslighting) as contextual factors disproportionately affecting this new patient population that further exacerbated their disability. Participants recognized that the seemingly invisible presentation of Long COVID led to disbelief and a lack of understanding from employers, insurance companies, family members, and healthcare professionals (HCPs). Stigma and testimonial injustice also resulted in increased financial disparities:

> Their employers and family physicians… don’t know why the patient is getting better and then suddenly not getting better and they are putting a lot of pressure on them.. they’re not… understanding what’s happening and they think the patients just don’t want to go back to work. (P13)

Participants described that some patients with Long COVID faced financial insecurity if unable to work. The inability to take time off work due to lack of access to disability supports exacerbated their symptoms and disability:

> If someone has the privilege of being able to take time off, of being able to focus on their recovery, [it’s] huge… people continue to get stuck in the Push-Crash [cycle] because they are trying to feed their family. (P10)

#### Pivoting to New Models of Physiotherapy Care Delivery

Although some participants absorbed patients with Long COVID into an existing model of care within their practice setting, many described how they pivoted to new models of physiotherapy care delivery (in-person to virtual models of care delivery; Long COVID clinics) as they encountered patients with Long COVID. Long COVID necessitated a shift from providing in-person to virtual care for patients living with Long COVID, recognizing that in-person visits drained patients’ limited energy levels and worsened Long COVID fatigue:

> It’s very challenging… to bring people in and do a therapy session because by the time they get dressed, drive, walk in from the parking lot, they’re exhausted. They have nothing left for me to do. I can do a couple things and then they go home and they are exhausted for the day so I am making them more sick rather than better. (P6)

Virtual care increased “*accessibility through the roof*” (P8), enabling flexibility and wider access to care (including to rural and remote settings). On the other hand, participants reported limitations with virtual care due to poor internet connection, unfamiliarity with technology, and difficulty creating meaningful engagement with patients over Zoom.

Additionally, many participants were introduced to this patient population through Long COVID-specific programs or models of care created to address the rising presentations of Long COVID. Though many physiotherapists in this study had little to no prior experience working with people with post-infectious illnesses, some described playing a key role in developing new programs of physiotherapy care, while others created treatment frameworks or updated courses to share knowledge among health care providers:

> About 4 months after the pandemic started…in July 2020 we had a request from our hospital to start a Long COVID clinic in our outpatient program. We didn’t really know anything about it– they were very clear that this was going to be the first program that we start. So, we’re gonna be learning kind of as we go. And we just sort of rolled with the punches. (P3)

### Learning Curves and Evolving Roles

Participants described their experiences of adapting and shifting their approaches as a cyclical and continuous process of learning curves and evolving roles, including: i) navigating uncertainty with Long COVID physiotherapy care; ii) keeping up with rapidly emerging evidence in Long COVID; iii) trial and error with Long COVID physiotherapy care by (a) drawing from past physiotherapy approaches used in other clinical populations, then b) learning from successes and failures in physiotherapy care), iv) adapting mindset and physiotherapy approaches to working with people living with Long COVID by (a) initially experiencing frustration and coming to terms with the limitations in physiotherapy scope, and b) re-evaluating expectations for goal setting and adapting physiotherapy approaches to ensure safe rehabilitation), which led to v) growing prominence of physiotherapy roles as advocates and collaborators fostering interprofessional partnerships in Long COVID care. (Figure 1).

#### Navigating Uncertainty with Long COVID Physiotherapy Care

Participants experienced uncertainty with Long COVID physiotherapy care, characterized by a lack of knowledge and understanding of the underlying mechanisms of this condition and the impact of their approaches to physiotherapy assessment and treatment:

> The only thing that we know is that the energy system is broken. We don’t know how to fix it…we don’t know prognosis, we don’t have any expectations, so the thing that’s really hard is that you have to recognize as a physiotherapist that you are limited in what kind of recovery you can offer. (P2)

Participants described their efforts to communicate their lack of knowledge and uncertainty with patients who also faced uncertainty regarding their long-term trajectory of Long COVID: “*I usually tell [patients] pretty upfront that…I don’t know it all*….” (P12)

#### Keeping Up with Rapidly Emerging Evidence in Long COVID

Physiotherapists acknowledged the importance of adhering to clinical practice guidelines, including the WHO guidelines for Long COVID rehabilitation (30). These guidelines provided a foundation that required tailoring to accommodate individual patient needs amidst the continually evolving evidence:

> It’s partly our responsibility as healthcare providers to be able to take that information that we have and then adapt it to the individual in front of us…to form kind of the best evidence treatment plan for the person in front of you. (P12)

However, participants discussed challenges and strategies regarding the abundance of emerging Long COVID research and clinical practice guidelines. Physiotherapists described limited time as a common obstacle in seeking out, appraising, and incorporating evidence into their practice:

> I think that it’s just the sheer amount [of readings…], and then having the time to be able to read through… I would say I don’t necessarily read a paper start to finish…with valid, trusted resources, you get a lot of good information. The struggle is fitting it in, because it can almost be a full-time job keeping up with it. (P2)

The rapidly evolving nature of Long COVID research presented difficulties for participants when trying to read, critically appraise, and interpret research evidence, due to changing guidelines and variations in the methodological quality of evidence regarding appropriate treatment. This made it difficult for participants to inform shared decision-making with their patients in relation to established clinical practice guidelines:

> The guidelines are new and ever-changing and there’s a ton of literature being pumped into this field, so I’ll have patients…and…clinicians bring me literature that conflicts with the guidelines…We’re still learning. (P8)

Although some participants mentioned their employers provided educational resources to support their professional development, such as newsletter updates, many noted an absence in training and took a proactive, self-directed approach to address knowledge gaps. They actively sought formal and informal learning opportunities to integrate into their practice including social media channels, podcasts, peer-reviewed literature, published guidelines, online webinars, websites, collaboration with interprofessional colleagues, and they attended conferences in their free time:

> …there was no training…even my onboarding to that job was basically…some websites you can look at, maybe connect with the concussion clinic or the complex chronic disease program that deals with ME/CFS [Myalgic Encephalomyelitis / Chronic Fatigue Syndrome], and so it was really kind of self, self learned…I basically figured that out. (P10)

#### Trial and Error with Long COVID Physiotherapy Care

Participants initially approached Long COVID rehabilitation by engaging in a process of “*trial and error*” that involved a) drawing on past physiotherapy approaches used in other clinical populations (including traditional exercise-based interventions, such as graded exercise therapy); resulting in b) learning from a series of successes and failures in physiotherapy care delivery.

##### a) Drawing on past physiotherapy approaches used with other clinical populations to Long COVID

Participants drew on their previous clinical experiences with other patient populations, noting similarities in symptoms between Long COVID and conditions such as acquired brain injury, chronic obstructive pulmonary disease, and ME/CFS. Two participants specifically highlighted drawing from their training in brain injury and concussion in addressing cognitive and sensory impairments in Long COVID:

> I work in brain injuries…and I remember thinking…what I am reading sounds a little bit like something [that] our post-concussion patients are reporting, not the same but there is some overlap there with some of the cognitive issues and the fatigue piece, and then…[the hospital directors] approached some of us on the brain injury team…[about] getting a COVID rehab program up and running. (P6)

Some physiotherapists relied on existing knowledge and training, applying traditional physiotherapy approaches including aerobic training and graded exercise therapy in Long COVID rehabilitation, under the assumption that patients were deconditioned, despite that these approaches may cause harm to people living with Long COVID with PESE or PEM. “*Fatigue is fatigue is fatigue. And when you treat it, the principles stay the same, you know, regardless of where it is coming from*” (P1). These physiotherapists applied exercise prescription as a universal approach across their patients living with Long COVID, rather than focusing on safe rehabilitation guidelines and biomedical evidence of physiological sources of fatigue highlighting impairments, highlighting gaps in knowledge about Long COVID and other infection-associated complex chronic illnesses:

> It was exercise prescription the way you would with any other patient who is deconditioned or struggling with respiratory symptoms… it was a lot like circuit training and education around aerobic and resistance training and getting them back to where they were pre-COVID. (P3)

##### b) Learning from Successes and Failures in Physiotherapy Care Delivery

Some participants strived to reflect and learn from their successes and failures, so they could better meet their patients’ needs. Many discussed how they adapted their approaches after recognizing the limitations of a deconditioning model. Namely, when applied to patients living with Long COVID, exercise resulted in symptom exacerbation and worsening of disability:

> So my initial assessments were just like every single myotome, head to toe, all range of motion…balance…a quick vestibular assessment… it was just horrendous…I put everybody through this and then they would crash…honestly my assessment now – we likely do very little. I want to find out…what their symptoms are, what they’ve been going through, what are they doing for physical activity, what is their social experience like. (P7)

Some participants distinguished between physiotherapy approaches best suited for post-intensive care unit (ICU) deconditioning versus Long COVID. One participant noted that patients with Long COVID often accessed the same rehabilitation programs as those with post-ICU deconditioning, posing a notable challenge for physiotherapists as they navigated distinct clinical presentations, recognized that their traditional approaches to deconditioning were harmful to Long COVID patients with PESE, and attempted to tailor their approaches accordingly:

> [Patients with Long COVID] were just crashing, they were trending in the wrong direction…we’re not helping… [But] there was a certain chunk of people [who]…had just come off a ventilator and got discharged home and they were on oxygen. Those people needed to exercise, but I wouldn’t classify them as a true kind of Long COVID, they were just post-ICU deconditioning. (P8)

We found variability across interview transcripts in participants’ level of knowledge and experiences working with patients with Long COVID as articulated by this participant: *“some physios are being overly aggressive, [not understanding] the level of fatigue they are dealing with…exercise is really contraindicated*.” (P6) Although many physiotherapists initially utilized a deconditioning approach which led to detrimental patient outcomes, they described evolving their approach to adapt safe rehabilitation and learn from their mistakes:

> Even myself, before I started my research, I thought Long COVID is…patients that are deconditioned…I have a lot of patients that before coming here, they went to other physios…and they were pushed a little bit too much, just a standard physiotherapy session. It is evolving and I think more and more people…are realizing the approach to it. (P13)

Twelve participants reported using virtual care with their patients. Participants were able to perform education, assessments, and functional tests (e.g., balance, gait, endurance). However, some described challenges overcoming some patients’ unfamiliarity with technology, poor internet connection, and no access to a webcam as well as difficulties establishing a meaningful rapport with their patients. In particular, observing patient progression via video-conferencing was challenging, but participants learned to optimize camera angles. As one participant described: *“[I made] sure that [patients] are visible on the screen and… they can see us.“* (P11)

#### Adapting Mindset and Physiotherapy Approaches

Participants described the need to adjust their approaches to physiotherapy working with people living with Long COVID, initially experiencing a) frustration and coming to terms with the limited scope of physiotherapy, and b) re-evaluating expectations for goal setting to ensure safe rehabilitation.

##### a) Frustration and Coming to Terms with the Limited Scope of Physiotherapy

Participants expressed frustration due to the unpredictable or limited success applying traditional physiotherapy approaches with persons living with Long COVID, followed by coming to terms with the understanding that physiotherapy interventions may not always yield improvements:

> …why isn’t someone who appears to be normal and who’s had regular testing by their physician, just not capable of improving their activity? It was frustrating at first to me ‘cause I didn’t understand why. But I also knew that there was something going on that would eventually kind of explain why that was happening. And I just had to be patient both with myself, and…my patients…as we came to understand [Long] COVID a little more and, and see what was going on behind the scenes and how it was impacting them. (P5)

Participants also described the role they played as a supporter, even if they were unable to offer certainty around treatment. Participants described their role as patient “*coaches*” and “*listeners*“:

Sometimes…it was just more listening and affirming that they were going through something major that was impacting their ability to do things like movement. (P5)

##### b) Re-evaluating Expectations for Goal-Setting and Adapting Physiotherapy Approaches to Ensure Safe Rehabilitation

Given limitations to the scope of physiotherapy and complexity of Long COVID, participants recounted importance of collaborative goal-setting; and in some cases the need to adjust their expectations to ensure supportive, accessible, and realistic approaches to rehabilitation with patients:

> I learned so much from Long COVID and my patients…through their lived experiences, their hospitalizations, that I had to completely change my idea of what a reasonable goal was. So I certainly had to temper my expectations for my patients…advocating for their return to independence but clearly understanding that that was not an achievable and realistic goal at the time, and having to change both of our expectations. Maintaining positivity, motivation and determination…but just framing it in a more accessible, realistic way. (P9)

Coupled with this change in expectations, participants described working towards more “*cautious*” (P11) approaches to rehabilitation that focused on preventing exacerbation of patients’ symptoms. Physiotherapists incorporated safe rehabilitation strategies such as pacing:

> We kind of have to be problem solvers…in terms of energy conservation…How do we break down these tasks for you so that you can actually get through activities of daily living [and which ones] are the most important to you? It’s a little bit of a different mindset…we’re used to really pushing people and getting them moving and exercising, and this is kind of the complete opposite, it’s coaching them to maybe do less and conserving energy and rest better…it’s primarily education and pacing management (P2)

Participants described attempts to employ rehabilitation approaches that minimized physical exertion in order to avoid symptom exacerbation, commonly utilizing breathing strategies, education, energy conservation, and fatigue management: *“The breathing pattern correction…when they’re breathing better, they can pace better…they can relax better*.” (P13)

#### Growing Prominence of Physiotherapy Roles in Long COVID Care

As participants progressed through their learning trajectories, many described embracing roles as a) Advocates: helping patients navigate services and mitigating stigma and discrimination, and b) Collaborators: fostering interprofessional partnerships in Long COVID care.

##### a) Advocate: Navigating Services and Mitigating Stigma and Discrimination in Long COVID Care

Participants described their role as an advocate for and with their patients when facilitating access to health services and by mitigating stigma and discrimination from other healthcare providers and employers in the context of Long COVID. They emphasized the importance of advocacy in helping patients navigate services, including assisting with insurance and supporting appropriate return to work. One participant educated patients’ employers about Long COVID rehabilitation by developing resources:

> We work closely with employers, we developed employer hand sheets to present. We were getting a lot of questions from employers saying, “Why is this person that looks normal still having symptoms that present like this so many months later?”…and we said, this is what you can expect with Long COVID, it’s not what you thought… (P5)

##### b) Collaborator: Fostering Interprofessional Partnerships in Long COVID Care

Participants described the importance of interdisciplinary collaboration with other providers such as occupational therapists (OTs), physicians, psychologists, and social workers to address the complexity of disability experienced by people with Long COVID. Participants emphasized that a multidisciplinary model “is a huge facilitator” (P4) for Long COVID care. One participant articulated that their patients who were accessing a multidisciplinary team “*get the most magnitude of improvement*” (P8) relative to those who were not, highlighting the importance of care that addresses all aspects of a person’s illness experience and symptomology. One participant described the different yet collaborative roles within the interprofessional team necessary to deliver comprehensive care:

> Very multidisciplinary, very interactive, highly collaborative because…physio was really thrown into Long COVID, …looking at breathing, we’re looking at function from a mobility perspective. But we’ve got people who can’t brush their teeth [or]…can’t do [some] more advanced [activities of daily living (ADLs)] that OT is well versed at…The OT was looking at mental health strategies and equipment and ADLs…Social work was looking at depression, suicidal ideation, counseling…Pharmacy would help patients navigate whatever medications [they’re] being prescribed and how they might interact with some…comorbidities… (P9)

A collaborative environment also facilitated the sharing of healthcare practitioners’ experiences including successes and failures, enabling participants to learn from each other’s expertise:

> I realized very, very quickly that a physio on their own might be helpful just to get the ball rolling in, in treating those patients [living with Long COVID]. But it wasn’t sufficient for the majority of those patients who had complicated issues. And so, understanding kind of, the impact [of disability] on their psychological health, [and understanding] their cognitive function helped kind of frame the treatment plan as well…And so…that approach was very different as opposed to treating a musculoskeletal problem or just a purely cardiovascular issue. (P5)

Overall, the experiences of physiotherapists in Canada working with adults with Long COVID in this sample involved a disruption requiring a shift in the profession and models of physiotherapy care delivery, followed by a cyclical journey involving learning curves and evolving roles of physiotherapists working with people living with Long COVID.

### Recommendations from Participants to Improve Rehabilitation for People Living with Long COVID

Participants identified recommendations spanning four areas at the individual, organizational, and health system level of rehabilitation care: i) integrate Long COVID-specific education and training into physiotherapy entry-to-practice level curricula and continuing education for current and future physiotherapists, including PESE or PEM screening and risks of harm with traditional exercise programs; ii) physiotherapists should engage as active and open-minded listeners with patients, iii) Long COVID rehabilitation needs to implement interdisciplinary models of care and learning to address the complexity of symptoms experienced by persons living with Long COVID, and iv) Organizational and system level improvements need to ensure publicly funded access to rehabilitation services in Canada, including addressing equitable access based on socioeconomic status, disability, race, ethnicity, gender, sexual orientation, immigration status, and other underserved groups. See Table 2 for each recommendation and supportive quotes.

**Table 2.**
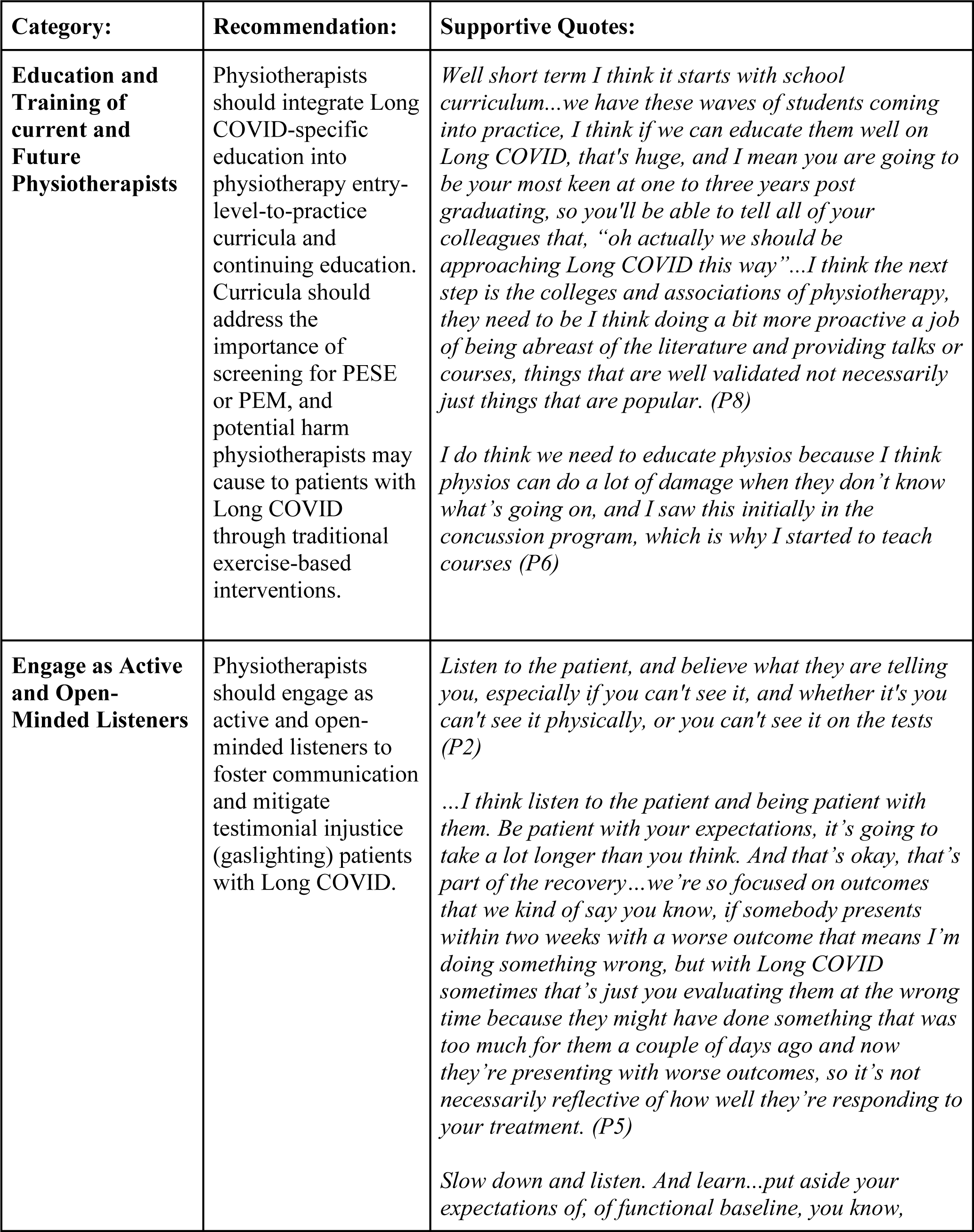
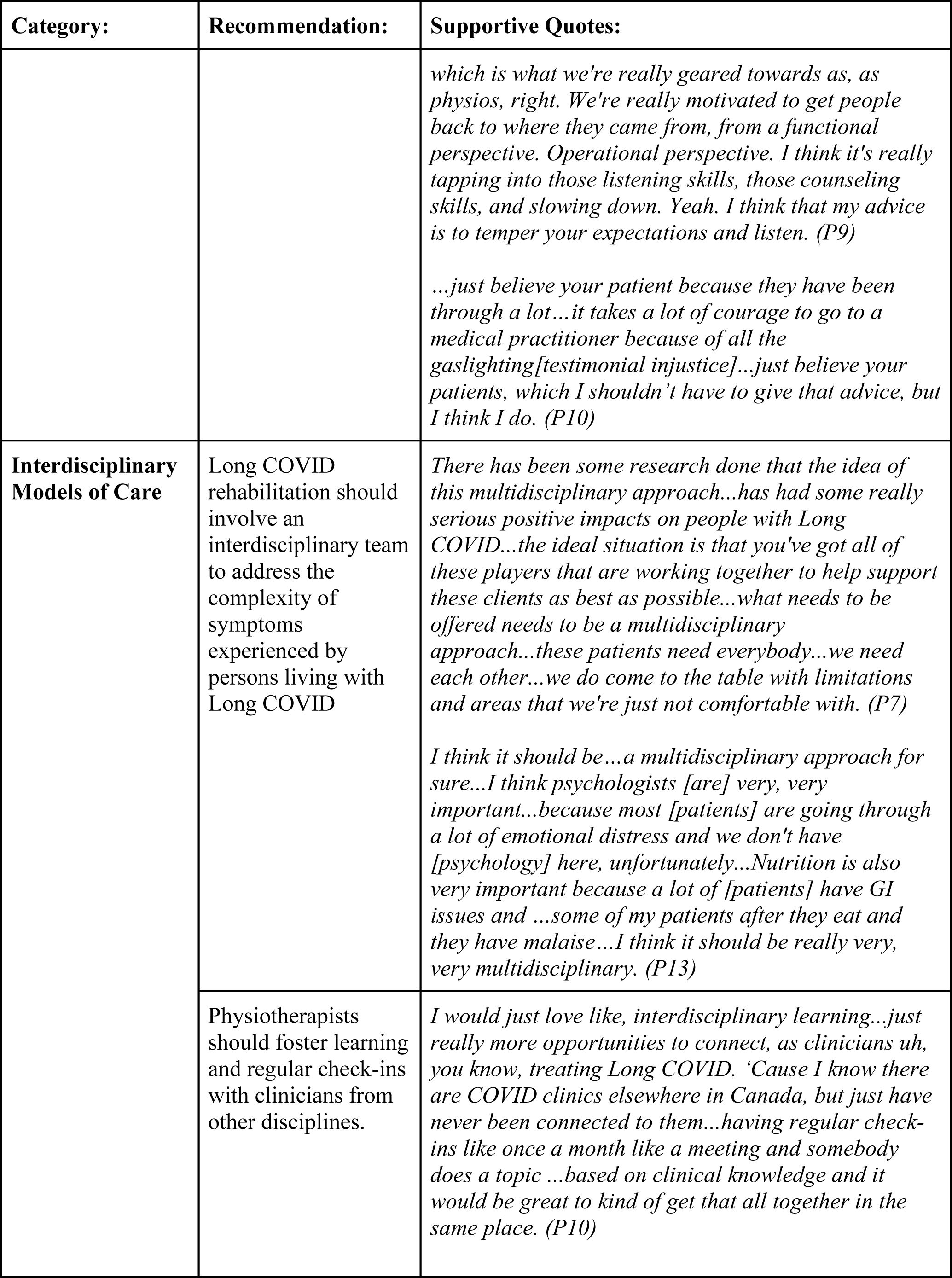
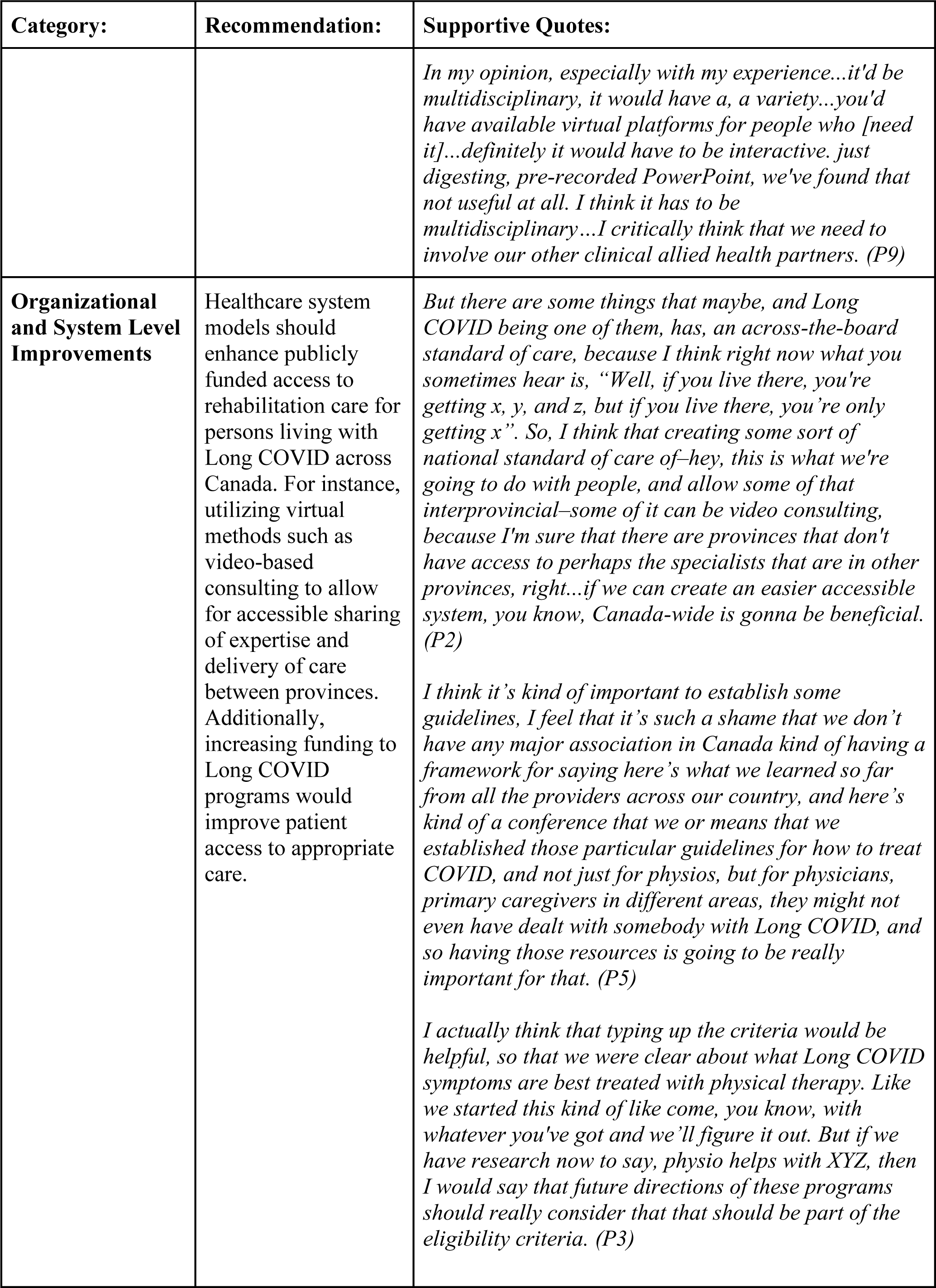
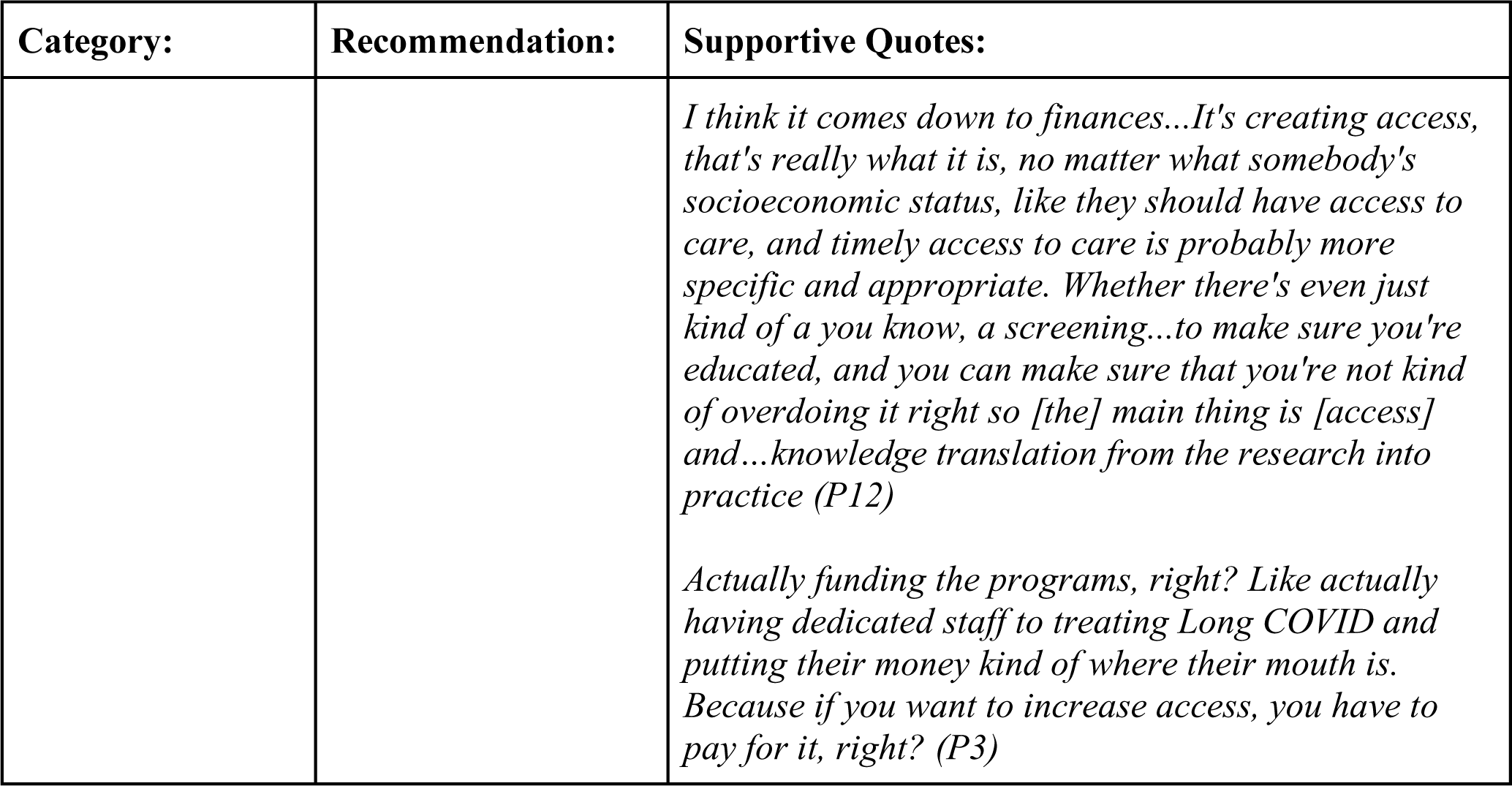
Recommendations from Participants to Improve Rehabilitation with People Living with Long COVID.

## DISCUSSION

Experiences of physiotherapists working with adults living with Long COVID were characterized as a disruption requiring a shift in the way they approached models of care delivery, giving rise to a dynamic process of learning curves and evolving roles as physiotherapists. Participants encountered a new patient population with complex multidimensional and episodic disability and pivoted to new models of physiotherapy care delivery delivered virtually and in Long COVID clinics. Physiotherapists responded by navigating learning curves keeping up with rapidly emerging Long COVID evidence and trial and error, navigating uncertainty, adapting mindset and physiotherapy approaches to Long COVID care, and adopting roles as advocates and collaborators working with persons living with Long COVID.

Throughout the cyclical learning journey, physiotherapists described taking on roles as a supporter, advocate, communicator, scholar, and interprofessional collaborator when working with patients living with Long COVID. These roles align with Canada’s Physiotherapy Core Competency Profile—Communicator, Leader, Collaborator, and Scholar (52). Despite variability in how participants drew upon these essential competencies within the context of Long COVID, these common roles emerged. In particular, the role of Communicator appeared prominent as participants emphasized the importance of transparency amid the uncertainties shared among physiotherapists and patients surrounding Long COVID recovery (53). Additionally, some physiotherapist participants appeared to embody roles as Leader and patient advocate as demonstrated by their descriptions of educating employers, insurers, families, and fellow healthcare providers and efforts to mitigate stigma and testimonial injustice experienced by patients (52,54). This holds particular significance given Long COVID’s disproportionate impact on those facing financial disparities and marginalized communities – especially racial, ethnic, and sexual and gender minorities (55–58). Furthermore, it was essential for physiotherapists to take on the role of supporter, as their patients navigated illness-related adjustments and identity loss (59,60).

However, participants in this sample highlighted the variability in clinical practice and approaches to working with adults living with Long COVID, along with differences in knowledge regarding guidelines and evidence-informed safe rehabilitation for this population. This was evidenced by physiotherapists who emphasized graded exercise and a deconditioning approach to rehabilitation (used with other populations), demonstrating their lack of understanding of existing guidelines on Long COVID rehabilitation (30). Given the similarities between Long COVID and other chronic and episodic conditions, such as ME/CFS, physiotherapists can gain valuable insights from the extensive evidence on precautions related to physical activity interventions in these conditions (12,61,62). Yet, some participants appeared to utilize traditional deconditioning approaches by ‘pushing exercise’ and prioritizing exercise progression without addressing PESE or PEM, and therefore risked patient harm (12,14,59,62). Some participants in this study appeared to neglect consideration of underlying sources of fatigue while favouring graded exercise therapies, leading to exacerbated disability for their patients living with Long COVID and causing a “crash” phenomenon (31). On the other hand, participants who described re-evaluating and adapting their approaches appeared to adopt existing guidelines on safe rehabilitation, minimizing harm and tailoring to the individual’s needs and challenges (30,31). Existing Long COVID clinical practice guidelines from the WHO that were available at the time of the interviews prioritize red-flag screening, individualized care, psychological support, and avoiding graded exercise therapy in the presence of PESE or PEM (30,31). Thus, it is critical for physiotherapists to remain up-to-date with evolving literature to ensure safe rehabilitation focusing on symptom stabilization and optimising function while preventing exacerbation of symptoms (30,32). There is a need for enhanced Long COVID education within physiotherapy entry-to-practice curricula and continuing education opportunities for physiotherapists to foster evidence-informed, safe, and effective rehabilitation with persons living with Long COVID. Our recommendations align similarly with conclusions drawn by a qualitative study regarding the experiences of healthcare providers in Long COVID rehabilitation conducted in Alberta whereby authors emphasized the importance of healthcare provider education to recognize the condition and current practices in Long COVID care, as well as interprofessional collaboration across practice settings (63).

### Strengths and Limitations and Implications for Future Research

This study employed a qualitative approach using semi-structured interviews, which allowed for in-depth exploration into perspectives of Canadian physiotherapists’ roles, experiences with assessment and treatment, knowledge acquisition, and facilitators and barriers influencing rehabilitation delivery in Long COVID care. Our team-based analytical approach and partnership with physiotherapists living with Long COVID enabled valuable collaboration, guidance, and advice for the refinement of the interview guide and demographic questionnaire, which increased the robustness of the study. Other strengths of this study included the variability in the number of patients living with Long COVID treated by each participant, as well as the diversity in participant characteristics across five Canadian provinces from three categories of clinical practice settings. Notably, few physiotherapists specialized in common post-infectious complex chronic conditions (eg. Long COVID, ME/CFS, Lyme disease), emphasizing the importance of interviewing individuals with varying levels of experience to provide a representative view of physiotherapists across Canada. Involving physiotherapists with lived and living experiences of Long COVID on the research team helped to enhance the quality of the interview guide, demographic questionnaire and offered guidance with our analysis. Our group-based approach to analyzing the interview transcripts helped to ensure rigour by incorporating perspectives across the research team.

This study reflects the perspectives of a small sample of physiotherapists who self-identified as working with people living with Long COVID, and does not reflect the lived experiences of people living with Long COVID. While it was beyond the scope of the study to include both physiotherapists and patients with Long COVID, it is important to interpret findings in conjunction with evidence representing experiences of people living with Long COVID accessing rehabilitation (60). Furthermore, the diversity of clinical experiences among physiotherapists reflected in the interview data suggests Long COVID clinics may have used variable definitions of Long COVID which resulted in some physiotherapists having experiences working with people who met the WHO definition of Long COVID and others working with people with symptoms from acute COVID-19 persisting less than two months or prior to three months from the onset of COVID-19 (3). Since participants primarily worked with adults and practiced in urban settings in Canada, these study findings may have limited transferability to physiotherapists working with children (who also experience Long COVID) (64,65), practising in other geographical settings, including rural or remote areas or working within different healthcare systems. Future research may explore the experiences of physiotherapists working with diverse patient populations, other countries and healthcare settings in the context of Long COVID. **CONCLUSION**

Experiences of physiotherapists were characterized by a dynamic process of a disruption to physiotherapy care delivery and a cyclical journey involving learning curves and evolving roles when working with individuals living with Long COVID. Results demonstrated the variability in knowledge and experiences among physiotherapists in this field, highlighting an urgent need for education among physiotherapists to ensure safe rehabilitation for people living with Long COVID.

## Data Availability

All data relevant to the study are included in the article or uploaded as supplementary information. Selected anonymized qualitative data from the interviews could be made available on request.

## Acknowledgements

This research was completed in partial fulfillment of the requirements for an MScPT degree at the University of Toronto. The study was supported by a catalyst grant from the Rehabilitation Science Research Network for COVID, Temerty Faculty of Medicine, University of Toronto. We thank the participants for their time and sharing their insights for this study. We thank Alyssa Minor, Mary Burke, and Kiera McDuff, for reviewing the interview guide and demographic questionnaire and for their feedback with a mock interview.

## Contributors

This research was completed in partial fulfillment of the requirements for an MScPT degree at the University of Toronto. KKO, SCC, DAB, SC, and KM provided guidance throughout all stages of the research including development of the research protocol, analysis, interpretation of results and manuscript development. KKO, SCC, DAB, SC, and KM supervised CK, CL, MW, SAJ, AG, and NK who developed the protocol, recruited participants, determined eligibility and obtained consent, conducted the interviews, analyzed the data, and drafted the manuscript in partial fulfilment of requirements for an MScPT degree at the University of Toronto. CK, CL, MW, SAJ, AG, and NK (MScPT students) developed skills in qualitative research methodology in course work on ethics, research strategies, literature review, study design, and protocol development. KKO, SCC, DAB, KM and AM reviewed and provided comments on earlier drafts of the manuscript. CK, CL, and MW led the revisions and response to co-author feedback on the manuscript. All steps were closely reviewed and guided by advisors (KKO and SCC). KKO is the senior responsible author and guarantor of the study. All authors read and approved the final manuscript.

## SUPPLEMENTAL FILES

Supplemental File 1 – Interview Guide

Supplemental File 2 – Demographic Questionnaire

Supplemental File 3 –Research Ethics Board Approval

Supplemental File 4 - Information and Consent Form

