## Supplemental File 1 for "Experiences of physiotherapists working with adults living with Long COVID in Canada: a qualitative study"

### Interview Guide

**Preamble:**

Thank you *[name of participant]* for agreeing to participate in this study. My name is *[name of interviewer]*, I am a student in the Physical Therapy program at the University of Toronto, and I will be conducting the interview today. As you may know, my study group and I are interested in your experiences, insights and perspectives working with adults living with Long COVID in Canada. We are conducting this research study to explore physical therapists' experiences providing rehabilitation and key factors to consider when working with individuals living with Long COVID. *[pause for any questions]*

Today's interview will take approximately 60 minutes. I will also be audio recording this interview and taking notes to help better understand and solidify the points that have been raised. Knowing this, would you like to continue with this interview?

Have you read, and are you in agreement with what is involved in the study? Would you be interested in receiving a copy of the study summary by email following the completion of the study?

**Do I have your consent to continue with the interview?**

I will be asking a series of questions about your clinical experience, knowledge and resources, and perspectives on working in Long COVID care.

The demographic questionnaire will be split into two parts, with the first questions asked at the beginning of the interview and few remaining questions asked at the end of the interview. I will record these on the demographic questionnaire (share screen via Zoom). Your responses to the questionnaire will help us describe the general characteristics of the participants in this study. There are no right or wrong answers. *[administer Part 1 of the Demographic Questionnaire with share screen on Zoom]*

Please feel free to interrupt me to ask questions or clarifications, skip questions, take a break, or stop the interview altogether.

Do you have any questions for me before we begin?

***start recording***

---

**SECTION 1: EXPERIENCE WORKING WITH ADULTS LIVING WITH LONG COVID**

Long COVID, also known as post COVID-19 condition, is defined as a condition that occurs 3 months from the onset of a probable or confirmed SARS CoV-2 infection, with symptoms that last for at least 2 months and cannot be explained by an alternative diagnosis."(WHO, 2021)

**PERCEIVED NEED AND ROLE OF PHYSICAL THERAPY IN LONG COVID**

1) How did you first learn about Long COVID?
2) What would you say your **general knowledge** is, or what do you know about:
    a) Long COVID?
        i) *Possible topics to probe include:*
            - Definition of Long COVID (aka what is it?)
            - Number of people living with?
            - Clinical pathophysiology / manifestation?
            - Types of disability associated with Long COVID (physical, cognitive, mental, social inclusion, daily activities)?
            - Episodic nature of disability?
    b) About long COVID rehabilitation?
3) What **role** do you think physical therapists have in Long COVID rehabilitation?
    a) *Possible topics to probe include:*
        - Impact of physical therapy on adults living with Long COVID
        - Person-centered care
        - Role of patient advocacy e.g. episodic nature of condition
        - Goal setting
        - Mitigating stigma, providing social support, preventing multimorbidity
4) What do you think the need is for physical therapy in Long COVID rehabilitation?
    a) As opposed to other healthcare professionals?
    b) Preventing or mitigating episodic disability:
        i) Potential to help address physical, cognitive, mental-emotional, and social health challenges (episodic disability)
        ii) Role of PT with pacing as a rehabilitation strategy? (stop, rest, pace)

**KNOWLEDGE, RESOURCES AND EXPERTISE**

There is a wealth of Long COVID research that has transpired in the past two and a half years, however, knowledge translation of evidence to practice is still emerging. In 2021, a survey administered to 255 Austrian physical therapists showed that only 11% reported feeling sufficiently informed regarding Long COVID rehabilitation (Scheiber et al., 2021). We would like to gather your perspectives on the knowledge and resources pertaining to Long COVID rehabilitation.

5) From your perspective, **how equipped do you feel physical therapists are** in assessing and treating adults living with Long COVID?

6) Have you had any **learning opportunities, formal education, or training** on Long COVID rehabilitation provided to you, whether through your organization or elsewhere?
    a) *Possible topics to probe include:*
        i) Support and training from your organization

ii) Training from workshops, events, conferences, online resources, community groups
iii) Experience with other conditions with an episodic nature or with similar symptoms such as post-exertional symptom exacerbation
iv) Anything specific on safe rehabilitation approaches with Long COVID? (e.g. pacing)

b) What type of education or training do you think is needed for PTs to equip them for working with adults living with Long COVID?
c) What topics in the field of Long COVID do you want to learn more about?

7) What **guidelines, literature or other resources** on Long COVID, **if any,** guides your practice?

a) *Possible examples:* webinars, articles, published or not published guidelines, pamphlets, podcasts, scholarly resources, presentations, and forums.
b) *Probe/leads to Question 9*

8) What challenges have you encountered with keeping up-to-date with Long COVID evidence?

a) *Possible topics to probe include:*
   i) *Can you describe some of the strategies you have used to overcome these challenges?*
   ii) *Were there any particular resources that have been helpful for you?*
   *iii)* How do you typically go about finding updated guidelines or research?
   iv) Guidelines that they follow:
      1) WHO clinical guidelines on management of COVID-19
      2) World Physiotherapy Briefing Paper
   v) Workshops, forums or educational events they attend
   vi)
b) *If the participant works for an organization:* Has the organization you work for provided any support for evidence-informed practice or guidelines?
   *i)* *Possible probes:*
   - Who was involved in the decision-making,
   - How did the organization facilitate practice,
   - Were there any barriers faced,
   - What could have been done differently,
   - Any support from outside of organization
a) *Probe/leads to Question 10*

9) From your experience, what are some **facilitators** that help integrate evidence-informed guidelines into practice?

a) *Possible topics to probe include:*
   *i)* At the organization level? At the clinician-level?

*ii)* Awareness of clinical guidelines for safe and effective Long COVID rehabilitation

10) From your experience, what are some **barriers** to integrating evidence-informed guidelines into practice?

a) *Probes:*

i) virtual rehab, access to appointments, time for appointments, caseload, environment (e.g. clinic, rehab hospital)

11) Given what you know about Long COVID now, what do you wish you knew about providing Long COVID rehabilitation from the beginning?

a) If opportune: Do you have any anecdotes you'd be willing to share about a patient with Long COVID you encountered where you did not know how to provide support, and how you navigated that situation?

**[ASK PARTICIPANT IF THEY WOULD LIKE A BIO-BREAK]**

**CLINICAL PRACTICES**

Our study focuses on Long COVID rehabilitation, particularly on physical therapy. We defined rehabilitation as "a set of interventions that work to optimize function in everyday activities, support people to recover or adjust, and enable participation in meaningful life roles".(Stucki et al. 2018)(WHO 2021).

12) Can you tell me about your clinical **experiences of rehabilitation with adults living with Long COVID** in your work setting?

a) *Possible topics to probe may include:*

- Can you describe the typical characteristics of the people with Long COVID that you encounter in your work setting?
  - Attitudes and beliefs about Long COVID, the episodic nature of the disease
- Can you describe the practice setting (environment) where you see patients with Long COVID and the resources (time, equipment, modalities) you use with this patient population?
- How does your practice setting (e.g. private practice, rehab, acute, etc.) impact the delivery of Long COVID rehabilitation?
- How long have you worked/been working in this setting?
- What other health care providers did you collaborate with, if at all?
- Community organizations involved with their practice setting

13) How did you become **involved in working** with individuals living with Long COVID?

a) *Possible topics to probe include:*

- ○ Level of preparedness
- ○ Experience with other chronic or episodic conditions, for example:
  - Myalgic Encephalomyelitis [ME]

- Chronic Fatigue Syndrome [CFS]
- Human Immunodeficiency Virus [HIV]

iii) Motivations to work in Long COVID rehabilitation

14) How does Long COVID rehabilitation **compare with rehabilitation for any other patient populations** you have worked with in the past?

a) If the participant has worked with individuals living with ME and CFS before, the probe for **similarities and distinctions** in terms of physical therapy assessment, treatment, and outcome measures with Long COVID (e.g. pacing).

15) Can you describe what an average physical therapy **assessment** would look like for your patients living with Long COVID?

a) *Possible topics to probe for:*
   i) Any systematic methods or approaches
   ii) Types of goals established
   iii) Baseline testing commonly used
   iv) Subjective factors/interview
   v) Are there any red-flags to assess / be aware of
   vi) Has the assessment evolved over time?
   vii) Use of technology to facilitate assessment

16) Can you describe what **treatments** or **interventions** you use with your patients living with Long COVID??

a) *Possible topics to probe include:*
   i) Types of interventions used (including home management) (e.g. pacing)
   ii) Timing of intervention;
   iii) Timeline and duration of intervention delivery
   iv) Referral process
   v) What types of goals are made
   vi) Follow-up after interventions, how have these changed over time
   vii) Use of technology to facilitate rehabilitation (e.g. to monitor heart rate, pacing)
   viii) Strategies to prevent or mitigate exacerbations or episodes of disability

b) To wrap up: Do you have anything further to add regarding treatments you used?

17) Can you describe some **successes** you've had with treatments for your patients with Long COVID?

18) Can you describe some **challenges** you've had with treatments for your patients with Long COVID?

19) What are some of your most commonly used **outcome measures** and how has that experience been for your patients living with Long COVID?

a) *Possible topics to probe for:* Technology for monitoring symptoms (eg. pulse oximetry, heart rate, self-reported questionnaires; objective performance assessments).

20) From your perspective, what have your patients living with Long COVID told you that has been particularly helpful about their physical therapy care?

21) In your perspective, what might an **ideal**, physical therapy-led rehabilitation program for Long COVID look like?

a) *Possible topics to probe:*
   i) How can the impact of rehabilitation be ideally tracked with measures?
   ii) Method of rehab delivery (e.g., virtual, in-person, or hybrid, type of practice setting, use of tele-rehabilitation, home care)
   iii) Use of technology to facilitate rehabilitation
   iv) Patient values and preferences
   v) Evidence-informed approaches
   vi) What aspects of **patient safety** (including but not limited to physical, emotional, and cultural safety) should ideally be considered in Long COVID rehab?
      1) PESE considered and monitored before, during and after assessments and treatments?
      2) Patient informed or knowledgeable about the implications of physical activity on PESE?
      3) Emotional safety: consideration for patients who have been subjected to medical gas lighting and minimization of symptoms
      4) Consideration of impacts of intervention on cognitive fatigue
      5) Consideration of method of intervention delivery: in-person, virtual, hybrid, and effects this has on PESE and cognitive fatigue.
   vii) Factors to consider regarding feasibility and sustainability of rehabilitation

#### <u>SECTION 2: FACTORS TO CONSIDER FOR IMPLEMENTATION OF REHABILITATION IN LONG COVID</u>

**[ASK PARTICIPANT IF THEY WOULD LIKE A BIO-BREAK]**

#### PERSONAL CONTEXTUAL FACTORS

22) Do you have any eligibility criteria that you use to treat adults living with Long COVID? If so, can you expand on any?

a) *Possible probes:*
   i) How are self-reported symptoms from patients considered?
   ii) How do you determine who will benefit from your services?
   iii) What do you think or believe about Long COVID from what you've seen and experienced thus far from your patients?

iv) Has your perception of Long COVID changed after treating patients living with the condition?

23) During your experiences working with this population, did you notice any commonly-held beliefs, attitudes or thoughts **from patients**? If so, what was the impact on rehab?

a) *Possible probes:*
   i) Medical gas lighting experienced by patients
   ii) Fear of not being believed or having symptoms taken seriously
   iii) Fear of uncertain nature of episodic disability

24) *Question only for participants who self-identify as living with Long COVID:* How has your experience living with Long COVID informed your delivery of Long COVID rehabilitation?

a) *Possible probes:*
   i) Ability to relate and connect with patients
   ii) Understanding of specific symptoms (ex. PESE, fatigue, cognitive symptoms)
   iii) Challenges associated with variability of clinical presentation

**EXTRINSIC CONTEXTUAL FACTORS**

25) Are there any social barriers or "stigma" that might affect patient participation in Long COVID rehab?

a) *Only for participants who are living with Long COVID or had Long COVID:*
   i) Is there any existing stigma that could affect your willingness to participate or engage in rehab?

26) In your experience, has an individual's income impacted their ability to participate in or seek out Long COVID rehabilitation?

27) In your experience, how has social support affected outcomes of individuals living with Long COVID?

a) *Probe:* Likelihood of seeking out Long COVID rehab

28) From your perspective, what are some **facilitators** to **delivering** Long COVID rehabilitation at your organization/work setting?

a) National strategy of Long COVID rehab care in Canada.
b) Federal / provincial / territorial funding

29) What were some **barriers** you experienced delivering Long COVID rehabilitation, and what, if any, were the strategies used to overcome them?

a) *Possible topic to probe for:*
   i) Accessibility to rehab (e.g., cost, presence of wait-lists),
   ii) Extrinsic factors (e.g., stigma, social support, income),
   iii) Intrinsic factors (e.g., living strategies),
   iv) Personal attributes (e.g., age, multimorbidity)

#### <u>SECTION 3: RECOMMENDATIONS</u>

At this point we are nearing the end of our interview. I would like to ask for some of your recommendations:

30) In your perspective, **what can be done to enhance rehabilitation for adults living with Long COVID?**

31) What might help to **address any knowledge gaps and learning needs** that physical therapists may have when working with adults living with Long COVID?

- a) *Possible topics to probe for:*
  - i) How to better educate on Long COVID-appropriate assessments, interventions, and patient education,
  - ii) Dealing with uncertainty

32) In your perspective, what might an **ideal**, physical therapy-led rehabilitation program for Long COVID look like at the system level?

- a) *Possible topics to probe for:*
  - i) Method of delivery (e.g., virtual, in-person, or hybrid, type of practice setting)
  - ii) Patient values and preferences
  - iii) Evidence-informed approaches

33) How do you think the **broader healthcare system** in Canada could **better support** the delivery of Long COVID rehab?

34) If you had **one piece of advice** to give to a physical therapist on their first day of working in Long COVID care, what would you give?

#### CLOSING QUESTIONS AND REMARKS

35) Is there anything else you would like to mention regarding Long COVID rehabilitation, or suggestions (for individuals living with Long COVID, physical therapists working in Long COVID)?

36) Are there any questions you have for me?

That brings us to the end of the interview section, and we'll be finishing with the second half of the demographic questionnaire. I will now stop recording. Thank you very much for participating in our study. We greatly appreciate you taking the time to share your experiences and perspectives with us.

*(Administer second half of demographic questionnaire via share screen)*

As a token of appreciation, you will receive an attached $40 electronic gift card in your email. Is there a type of gift card you would prefer to receive? (Name the options we will be providing).
