## Supplemental File 2 for "Experiences of physiotherapists working with adults living with Long COVID in Canada: a qualitative study"

### Demographic Questionnaire

**[to be administered via a Word document with shared screen on Zoom – interviewer will enter responses into the Word document directly on screen after the interview]**

Thank you for taking part in the Long COVID and Rehabilitation study with the goal of examining the experiences of physical therapists working with adults living with Long COVID in Canada.

---

Thank you for completing the interview. This part of the study involves completing a 2-minute demographic questionnaire. Questionnaire responses will be grouped together to help us describe, in general, the characteristics of the participants who took part in the study and will not be linked to you as an individual.

**PERSONAL DEMOGRAPHIC QUESTIONS (the interviewer will highlight the responses directly in the Word document and will save the document with the P number.**

**Participant Number: [enter P number here]**
**Interview Date: [enter date here]**

**PART 1 - BEFORE THE INTERVIEW – Demographic Questions**

1) How many **years** have you been working as a physical therapist? ____________ years

2) How many **months** have you been working with adults living with Long COVID? _______ months

3) What **practice setting** do you currently work in? (highlight one response)
   a) Acute care hospital
   b) Rehabilitation hospital (private, work-related injuries)
   c) Private practice
   d) Home care
   e) Long term care
   f) Other (specify): ___________

4) What is the **city or town** that you currently practice in? _________________

5) What **province or territory** do you practice in? ______________________

6) Approximately **how many patients** have you treated in the past year that are/were living with Long COVID? An estimate is fine. ___________ patients.

7) Have you personally **had COVID-19** in the past? (highlight one answer)

a) Yes
b) No
c) Unsure

8) Do you have personal **lived experiences** of living with Long COVID? (highlight one answer)

a) Yes
b) No
c) Unsure

---

**PART 2 - AFTER THE INTERVIEW – Demographic Questions (highlight the response in yellow)**

| Question | Response options |
| --- | --- |
| 9) What is your **age** (in years)? | ________________ years |
| 10) What **sex** were you assigned at birth/ is on your birth certificate? | • Male<br>• Female<br>• Intersex<br>• Not listed (please specify): _____________<br>• Prefer not to answer |
| 11) What **gender** do you identify with? | • Man<br>• Woman<br>• Non-binary<br>• Prefer to self-describe (please specify): ______________<br>• Prefer not to answer |
| 12) Which **ethnicity** do you identify the most with? | • First Nations<br>• Inuit<br>• Métis |

| **Question** | **Response options** |
|---|---|
| | • Other Indigenous Peoples (please specify):_______________<br>• Asian<br>• Black or African<br>• Caribbean<br>• Native Hawaiian or Pacific Islander<br>• Hispanic<br>• White<br>• Not listed (please specify):___________<br>• Prefer not to answer |

**PROFESSIONAL DEMOGRAPHIC QUESTIONS**

| **Question** | **Response options** |
|---|---|
| 13) Which of the following best describes your **employment**? | • Employed - Full time PT<br>• Employed - Part time PT |
| 14) Have you worked with **children or youth** (<18 years old) in the past year who are living with Long COVID? | • Yes<br>• No |
| 15) Which **area of practice** do you most identify as currently working with? (check one that best applies) | • Musculoskeletal/orthopaedics<br>• Neurological<br>• Cardiorespiratory<br>• Paediatric<br>• Geriatric<br>• Leadership/Administrative/Management<br>• Research<br>• Other or Mixed (please specify): __________ |

| **Question** | **Response options** |
|---|---|
| 16) What **percentage** of your caseload is delivered virtually/remotely? (please provide a rough best estimate) | • 0% virtual<br>• 25% virtual; 75% in-person<br>• 50% virtual; 50% in-person<br>• 75% virtual; 25% in-person<br>• 100% virtual<br>• Other (please specify): ________________ |
| 17) Have patients <u>**accessed**</u> your physical therapy services specifically for Long COVID rehabilitation? | • Yes<br>• No<br>• Don't know |
| 18) Have you <u>**received physician referrals**</u> for any of your patients specifically for Long COVID physical therapy? | • Yes<br>• No<br>• Don't know |

**Thank you and closing remarks**

Thank you for taking the time to complete the questionnaire for our study examining the experiences of physical therapists working with adults living with Long COVID in Canada.
