## Supplemental File 3 for "Experiences of physiotherapists working with adults living with Long COVID in Canada: a qualitative study"

RIS Protocol Number: 43692

Approval Date: 24-Nov-22

PI Name: Dr Kelly O'Brien

Division Name:

Dear Dr Kelly O'Brien:

Re: Your research protocol application entitled, "Long COVID and Rehabilitation: Experiences of physical therapists working with adults living with Long COVID in Canada: a qualitative study"

The Health Sciences REB has conducted a Delegated review of your application and has granted approval to the attached protocol for the period 2022-11-24 to 2023-11-23.

This approval covers the ethical acceptability of the human research activity; please ensure that all other approvals required to conduct your research are obtained prior to commencing the activity.

Please be reminded of the following points:

- An **Amendment** must be submitted to the REB for any proposed changes to the approved protocol. The amended protocol must be reviewed and approved by the REB prior to implementation of the changes.
- An annual **Renewal** must be submitted for ongoing research. Renewals should be submitted between 15 and 30 days prior to the current expiry date.
- A **Protocol Deviation Report** (PDR) should be submitted when there is any departure from the REB-approved ethics review application form that has occurred without prior approval from the REB (e.g., changes to the study procedures, consent process, data protection measures). The submission of this form does not necessarily indicate wrong-doing; however follow-up procedures may be required.
- An **Adverse Events Report (AER)** must be submitted when adverse or unanticipated events occur to participants in the course of the research process.
- A **Protocol Completion Report** (PCR) is required when research using the protocol has been completed.
- If your research is funded by a third party, please contact the assigned Research Funding Officer in Research Services to ensure that your funds are released.

Best wishes for the successful completion of your research.



**OFFICE OF RESEARCH ETHICS**
McMurrich Building, 12 Queen's Park Crescent West, 2nd Floor, Toronto, ON M5S 1S8 Canada
Tel: +1 416 946-3273 ● Fax: +1 416 946-5763 ● ● http://www.research.utoronto.ca/for-researchers-administrators/ethics
