## Supplemental File 4 for "Experiences of physiotherapists working with adults living with Long COVID in Canada: a qualitative study"

**INFORMATION SHEET & CONSENT FORM**

**MScPT Student Researchers:**
Shahd Al Hamour Al Jarad, BHSc (Hon), MScPT[1]
Amy Gao, BHSc (Hon), MScPT[1]
Nicole Kaufman, BHSc (Hon), MScPT[1]
Caleb Kim, BHSc (Hon), MScPT[1]
Chantal Lin, BSc (Hon), MScPT[1]
Michelle Wong, BHSc (Hon), MScPT[1]

**Co-Principal Investigator (University of Toronto)**
Dr Kelly O'Brien, BScPT, PhD
Associate Professor
Department of Physical Therapy University of Toronto 500 University Avenue, Room 160
Toronto ON

(416) 946-3935

**Co-Investigators**
Dr. Soo Chan Carusone, PhD
McMaster Collaborative for Health and Aging
McMaster University
Hamilton, Ontario, Canada


Darren Brown, PT, FCSP, MSc, Mres
Long COVID Physio
Chelsea and Westminster Hospital, NHS Foundation Trust
London, United Kingdom


Dr. Saul Cobbing, PT, PhD
University of Toronto, Toronto, Ontario, Canada
University of KwaZulu-Natal, Durban, South Africa


Kiera McDuff, PT
Program Coordinator, Rehabilitation Science Research Network for COVID
University of Toronto, Toronto, Ontario, Canada


**SPONSORSHIP**
This study is supported by the Rehabilitation Science Research Network for COVID, Temerty Faculty of Medicine, University of Toronto

**INTRODUCTION**
You are being asked to participate in a research study, which aims to describe the perspectives of the experiences of physical therapists working with adults with Long COVID in Canada. We will use one-on-one interviews to explore your clinical experiences in Long COVID, knowledge and resources, perspectives on the role, implementation, and evaluation of rehabilitation, and lastly, and recommendations for future practice. The research team consists of six physiotherapy students from the University of Toronto, who are under the supervision of Dr. Kelly O'Brien (Associate Professor, University of Toronto) and Dr. Soo Chan Carusone (Director, McMaster University), Darren Brown (Chair, Long COVID Physio), Dr. Saul Cobbing (Visiting Professor, University of Toronto), and Kiera McDuff (Coordinator, University of Toronto). This study is a collaboration with Long COVID Physio.

Please take your time to ask any questions you have regarding your participation in our study. You will be asked to provide verbal consent at the beginning of the interview, which will be documented by the researcher at the end of this form. Providing your consent to participate in this study is entirely voluntary and declining our request to participate will not affect your relationship with the researchers (if applicable).

**WHY IS THIS STUDY BEING DONE?**
Given the growing prevalence and multi-dimensional nature of disability caused by Long COVID, rehabilitation is well-positioned to address the negative impacts experienced by individuals living with this condition. Physical therapists are at the forefront of preventing, mitigating, or addressing disability among individuals living with Long COVID. However, the experiences of Canadian-based physical therapists working with individuals living with Long COVID, specifically perceptions of the role, knowledge, and clinical practice are unknown.

**WHAT IS THE PURPOSE OF THIS STUDY?**
The purpose of this research study is to explore experiences working with adults with Long COVID from the perspectives of physical therapists in Canada.

**WHO CAN PARTICIPATE IN THIS STUDY?**
You are able to take part in this study if you:
- Are a physical therapist registered to practice in Canada <u>and</u>
- Have clinical experience within the past year working with adults living with Long COVID:

Clinical experience can be in any setting (does not have to be a Long COVID designated clinic), and may involve assessment and treatment of 1≥ adult(s) living with Long COVID within a larger caseload.

You will require access to a computer (desktop or laptop), tablet or smart phone with access to the internet. You have the option of either downloading the Zoom application (free of charge from: https://zoom.us/download) or launching Zoom from your web browser. In order to participate in the interview using Zoom, you will require the use of a microphone. We also recommend the use of headphones and a webcam; however, these are optional.

**WHAT IS INVOLVED IF I PARTICIPATE?**

If you are eligible, interested and agree to participate in this study, you will be asked to take part in an interview with two student researchers and complete a demographic questionnaire. The interview will be conducted online using the teleconferencing platform, Zoom (with the camera feature on Zoom preferred, but not required).

During the interview, we will ask a series of questions about your perspectives and experiences of working with people living with Long COVID. We are interested in your perspectives and experiences providing Long COVID rehabilitation, the need for physical therapy for this population, different factors to consider when working with the Long COVID population, and recommendations for future clinical practice.

The interview will be audio recorded (using a separate audio recorder, not on Zoom), typed out word for word at a later date and notes will be taken throughout the interview. If you are not comfortable answering any questions, you may choose to skip questions, take a break, or end the interview at any point with the option to reschedule.

After the interview, you will be asked to complete a demographic questionnaire (word document shared on the screen where we will enter the responses directly) that asks questions about your age, gender and length of time and type of area of your physical therapy practice. The total estimated time to complete the questionnaire and interview is 60-90 minutes.

**HOW WILL MY INFORMATION BE KEPT PRIVATE?**

All information you provide throughout the study, including demographic information, the consent form, notes throughout the interview, audio recordings, and transcripts will be only accessible by the research team and members of the University of Toronto Health Sciences Research Ethics Board, who have reviewed the study. A participant number will be assigned to you for identification, hence your name will not be used nor disclosed throughout the entire study.

We will use Zoom (version 5.11) Best Practices guidelines for the interview: The meeting room will be locked with a password (provided to you in advance of the interview), interviewers will manually accept you from a secure waiting room, and we will use a separate audio recorder to optimize privacy during the interview. We recommend you find a quiet, private area for participating in this interview.

All audio recordings from the interview will be transferred to ShareFile, a secure, encrypted data storage platform provided by the University of Toronto immediately following the interview, and the files on the recording device will be deleted. The researchers will complete the interviews online via Zoom in a private, secure, and quiet space at either the interviewer's private residence, internship locations (e.g: clinics, rehabilitation centers or hospitals), or the Episodic Disability and Rehabilitation lab at the University of Toronto (500 University Avenue). The audio files and electronic files (questionnaire responses) will then be downloaded to a secure, password protected folder at the University of Toronto. Your name will not be on these files (only your participant number). The audio recordings will be typed out word for word following each interview by the same MScPT students involved in the original interview.

**WHAT ARE THE POSSIBLE RISKS?**
There are no known physical or emotional risks of participating in this study, however it may be possible that certain questions may be uncomfortable for you to answer. As a result, you may choose to either skip questions or not answer them, or end the interview at any point.

**WHAT ARE THE POSSIBLE BENEFITS?**
There are no direct benefits for participants in taking part in this study. However, the knowledge gained from your perspectives will inform future research and practice, which may in turn improve physical therapy for adults living with Long COVID.

**WILL I BE PAID TO PARTICIPATE IN THIS STUDY?**
You will not be paid to take part in this study; however, you will receive a $40 CDN electronic gift card (E-gift card) as a token of appreciation for your participation in the study. If you choose to withdraw from the study during the interview, you will still receive the E-gift card.

**WILL THERE BE ANY COSTS?**
Your participation in this research project will not involve any additional costs to you. You will require access to a computer (desktop or laptop), tablet or smart phone with access to the internet. You have the option of either downloading the Zoom application (free of charge from: https://zoom.us/download) or launching Zoom from your web browser. In order to participate in the interview using Zoom, you will require the use of a microphone. We also recommend the use of headphones and a webcam, however these are optional. Zoom also includes many security features that will allow your information to be kept safe (described above in the "How Will My Information be Kept Secure?" section).

**WHAT WILL HAPPEN TO THE INFORMATION FROM THIS STUDY?**
We will submit a manuscript for publication and present a poster at the University of Toronto, Department of Physical Therapy Research Day upon the completion of the study in Summer 2023. If you are interested in receiving information from this study, please indicate this on the consent form.

**WHAT IF I DO NOT WANT TO TAKE PART IN THE STUDY?**
You are free to decide if you want to take part in this study or not. If you decide not to take part, or you withdraw at any time, it will not affect your relationship with the researchers (if applicable). If you decide to withdraw from the study, the research team may use the information collected from the interview unless you request your data to be destroyed. You will have up to 7 days after the completion of the interview to request that the data collected from your interview not be used and destroyed.

**IF I HAVE ANY QUESTIONS OR CONCERNS, WHOM CAN I CONTACT?**

**CONTACT INFORMATION**

If you wish to participate in this study, you may contact the student researchers at or call (416) 946-3935.

**CONSENT TO PARTICIPATE**

**Experiences of physiotherapists working with adults living with Long COVID in Canada: a qualitative study**

I have read and understood all of the content of the "Information Sheet". In no way does signing this form waive my legal rights nor relieve the investigator and sponsors from their legal and professional responsibilities.

By signing or giving verbal consent below I am agreeing that:

❑ I understand the information provided for the above study

❑ I have been able to consider the information, ask questions, and have had them answered thoroughly

❑ I understand that my participation is voluntary and that I am able to withdraw my consent for this study up to 7 days after the completion of the interview **without** any penalty.

❑ I understand that the data collected during the study may be looked at by individuals from the research team, or the regulatory authorities where it is relevant

❑ I give permission to these individuals to have access to the information I provided

❑ I give permission to allow the University of Toronto students to conduct this interview.

❑ I agree to participate in the above study

❑ **Provision of written consent**

____________________________ Participant's Name (please print)

____________________________ Participant's Signature

___________________________ Date

❑ **Provision of verbal consent**

______________________________ Researcher's Name (please print)

______________________________ Researcher's Signature

________________________________ Date

I would like to receive a **copy of the study summary** by email following the completion of the study.

❑ Yes, Contact:_______________________________________________
❑ No

I would like to be **contacted** in the case of **future research** looking at research about Long COVID and rehabilitation.

❑ Yes, Contact:_______________________________________________
❑ No

I, the undersigned, have fully explained the study to the above participant.

Researcher's Name (please print) ___________________ Date _____________________

Researcher's Name (signature) ____________________ Date ______________________
